# Examining Effective Patient-Provider Communication in Antenatal Settings across England: An In-Depth Analysis of Woman’s Experiences

**DOI:** 10.1101/2024.05.02.24306775

**Authors:** Martyna Andrzejczak, Gareth Elfed Jones, Gareth Anthony Nye

## Abstract

Pregnancy requires appropriate guidance and support from healthcare professionals. Understanding women’s experiences of effective patient-provider communication through antenatal care is critical, as evidence suggests, inadequate communication could pose various risks to maternal and neonatal well-being. Despite efforts to modernise maternity services, recent assessments reveal persistent challenges, with nearly half of maternity services inspections categorised as ‘inadequate’ or ‘requires improvement’. This qualitative systematic review investigated effective patient-provider communication in antenatal care settings across England, contributing essential insights into women’s experiences and feelings from various backgrounds.

**Methods:** This study utilised the PICo framework to formulate the research question, focusing on effective patient-provider communication in antenatal care settings across England. A comprehensive search involving various study types was conducted across electronic databases from 2010 onwards. Inclusion and exclusion criteria were predefined, and relevant studies underwent thorough screening. Data synthesis involved a qualitative descriptive approach, employing thematic analysis to capture diverse experiences. Findings were summarised through coded extracts, supporting quotes, and a narrative addressing women’s experiences.

**Results:** The search identified 46 records, with six studies meeting the inclusion criteria. Results were synthesised utilising thematic analysis approach. Subsequently, five themes were distinguished: *Responsive and engaging communication; Individualised treatment; Clear presentation of service information and informed choice;* Continuity *of care*; and ‘*Additional ways of communication’.* The study identified key suggestions to improve patient-provider communication in antenatal care, including tailored training programmes aimed at HCPs focusing on empathy, active listening, building emotional connections, establishing trust, and providing continuity of care. Recommendations also emphasise transparent information and empowering women through communication. Likewise, suggestions extend to the incorporation of cultural safety training initiatives and addressing structural issues within the system. The study, however featured experiences of minority ethnic women, which may potentially impact results, limiting the findings generalisability. In addition, measuring women’s views amid the emotional intensity associated with pregnancy presents inherent challenges, hypothetically affecting the depth of understanding of the experiences.

**Conclusion:** Few key conclusions derived from women experiences highlight the need to re-address barriers to communication. Proposed strategies offer practical steps, but further research is urged to address emerging challenges by advocating for improved communication strategies in order to enhance prenatal care nationally.

## Introduction

### Contextual

Antenatal care services in England are offered free of charge by the National Health Service (NHS), and they facilitate an approach adapted to the varying risk profiles of pregnant women. The following system structure ensures that low-risk pregnancies receive midwifery-led care, while high-risk cases receive additional care from obstetricians and consultants (NICE, 2021). Antenatal stage also assists foetal ontogenetic development and is an exceptionally challenging period in which women are exposed to major changes to their anatomy, physiology, and emotional state. Similarly, this time is commonly indicative of the lengthiest phase of healthcare engagement that women undergo at that point in their lives. There are some challenges in this setting, particularly as both the mother and the developing baby require care and attention, meaning a substantially broader group of healthcare professionals (HCPs) could be involved in their care, highlighting the importance of adequate patient-provider communication. Throughout this time, effective communication between healthcare providers and pregnant women is critical in ensuring the well-being of the mother and the unborn child (Watson et al., 2016). This way of communication is defined as the process wherein the sender articulates their message with precision and clarity, ensuring the receiver accurately comprehends the intended message, which results in a mutual exchange that establishes a shared and satisfactory understanding between both parties involved (Ratna, 2019) (see **Figure 1**).

**Figure 1.**
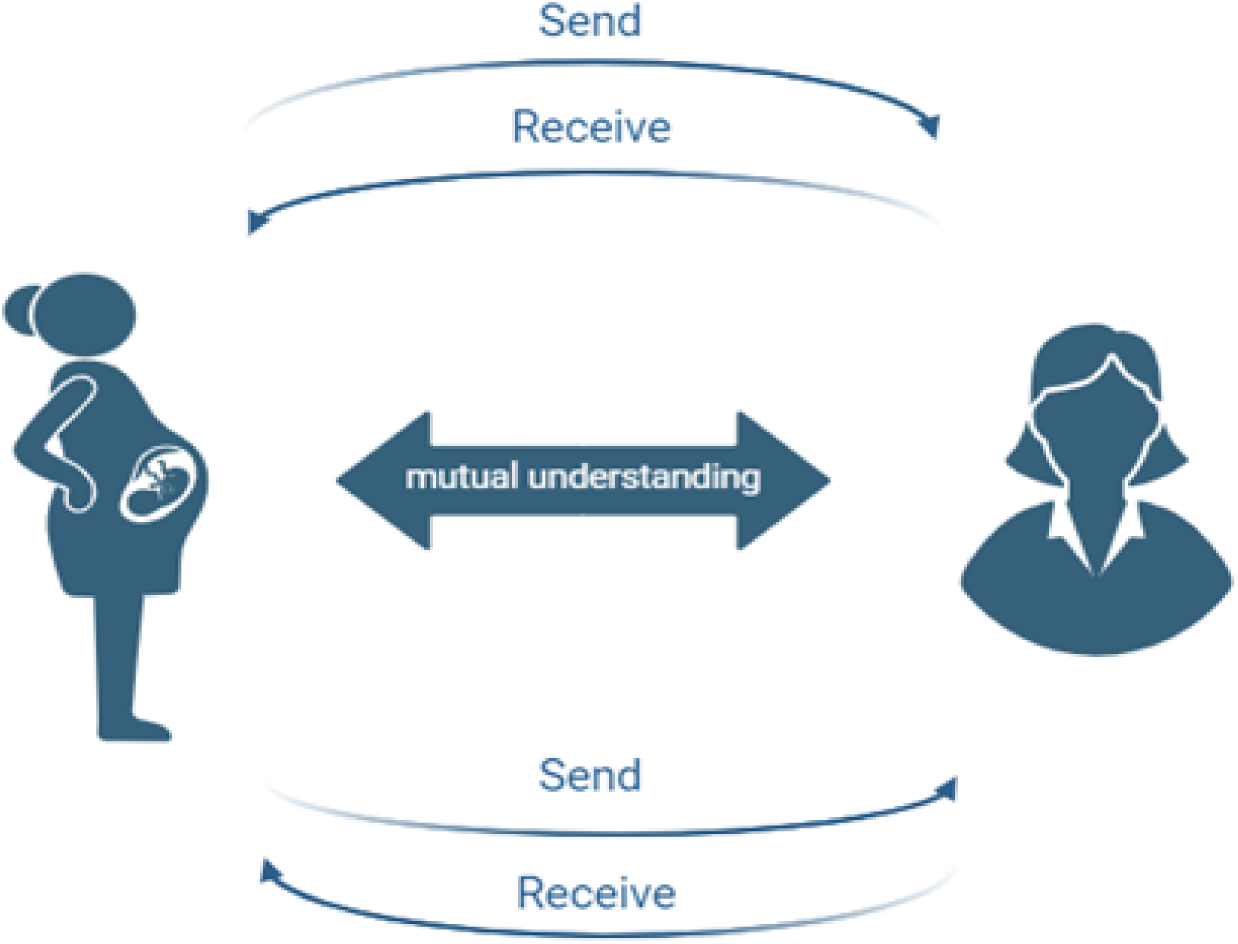
Effective patient-provider communication. Figure created with BioRender.com, illustrates a process of effective patient-provider communication in antenatal care settings aimed at ensuring shared understanding as described by Ratna (2019).

### Significance of Effective Patient-Provider Communication

According to Vamos et al. (2019) negative experiences with communication during pregnancy can significantly influence a woman’s emotional well-being and overall health, both in the short term and over an extended period. A qualitative study by Rowe et al. (2002) demonstrates that effective interactions are critical for patient satisfaction. Stating that pregnant women sincerely rely on the support delivered by their providers during prenatal appointments, and that they appreciate clear and well-executed communication between themselves and the providers. Additionally, they value involvement in decision-making concerning their care and addressing their questions and worries. It is thought that the involvement shown by healthcare professionals through appropriate communication during antenatal care creates a feeling of safety, trustworthiness, and reassurance, which are crucial for the welfare of the mother and her baby (Fraser, 1999). Regrettably, the healthcare system does not consistently provide the care and attention women require through this transformative journey. In concordance with Nicoloro-SantaBarbara et al. (2017), patient-provider communication difficulties have been identified as one of the factors of poor obstetric outcomes. The findings of this study are supported by the Schmiedhofer et al. (2021) report, which states that communication is indeed a barrier to effective and safe prenatal care. In England, this matter already became apparent in 1993, when the Expert Maternity Group established by The Department of Health (DH) published a report titled ‘Changing Childbirth’, which reviewed maternity services in England, stressing the magnitude of appropriate communication between medical personnel and expectant mothers (DH, 1993). Since then, increasing evidence has highlighted the importance of good-quality interactions between patients and carers in prenatal settings. Notably, in 2008, The Kings Fund’s independent inquiry identified data on maternal mortality in the United Kingdom from numerous studies and found insufficient communication as a factor contributing to it (O’Neill, 2008).

Moreover, Chen et al. (2007) concluded that inadequate antenatal care related to poor neonatal outcomes, where pregnancies were complicated by relatively common conditions such as anaemia, diabetes, or hypertension. Other studies also suggest that negative experiences attributed to poor communication during pregnancy care can equally have damaging implications for the foetus, which is specifically sensitive to environmental disruptions from maternal encounters, including exposure to psychological and biological stressors (Coussons-Read, 2013; Monk et al., 2012). Importantly, Rowe et al. (2001) and De Bernis et al. (2016) also emphasised the significance of adequate prenatal care and reported that poor communication may contribute to stillbirths and infant deaths. Although the significance of effective communication between expectant mothers and healthcare providers during antenatal care (ANC) in England has been acknowledged and recognised, as demonstrated by various initiatives such as the 6Cs framework (DH, 2012), the Five Year Forward View (NHS, 2014), and Better Births (National Maternity Review, 2016), all aiming to provide modernised maternity services, accessible information, utilise appropriate language, emphasise patient-centric approaches, and uphold NHS principles, with patient being at the heart of all interferences.

### Research Gap and Study Objectives

Still, it is concerning that despite these measures, nearly half of the inspections conducted by the Care Quality Commission (CQC) on maternity services in the UK result in safety evaluations categorised as either ‘inadequate’ or ‘requires improvement’ (CQC, 2018). Moreover, the most recent assessments, such as the National Maternity Survey 2022, also conducted by the CQC, indicate persistent challenges (CQC, 2022). A concerning trend in the survey suggests that maternity services in England are falling short of women’s expectations. This decline is marked by a decrease in women’s confidence in staff, with nearly one-third needing more trust and confidence in staff delivering their care. This trend is not unique to England and is seen globally, where substandard care in various maternity services contributes to preventable maternal and neonatal mortality and morbidity (WHO, 2017). It is stressed that every pregnant woman is entitled to high-quality, person-centred, safe, and effective healthcare (Crowe & Manley, 2019). Moreover, studies worldwide have highlighted the importance of appropriate patient-provider communication and its impacts on pregnancy care (Knapp et al., 2011; Kozhimannil et al., 2015; Lerman et al., 2007). However, the lack of synthesised data on women’s experiences engaging and interacting with healthcare providers through ANC in England is a concerning gap in the literature. It is, therefore, crucial to conduct an examination of the experiences of pregnant women in England from diverse backgrounds considering these challenges.

A comprehensive exploration is vital for understanding the complex issues related to communication throughout ANC. It is hypothesised that improved communication between healthcare providers and expectant mothers leads to better prenatal care experiences, higher levels of trust and confidence in healthcare staff, and ultimately contributes to enhanced maternal and neonatal outcomes. Addressing and enhancing the quality of prenatal care in the context of effective patient-provider communication on a national level stands as a priority. The aim of this study is thus to provide synthesised insights that contribute to the advancement of current knowledge on this topic and to explore women’s experiences in England during the antenatal period, focusing on effective patient-provider communication. Moreover, the study seeks to understand the emotional impact of challenges on expectant mothers and their satisfaction with prenatal care while identifying key issues they face. By synthesising qualitative studies, investigating common themes across experiences and challenges in women’s interactions with healthcare providers and assessing their impact, this research offers insights into improving prenatal care practices and enhancing the well-being of mothers and their children in England.

## Methods

### Study Questions/Literature Search Strategy

The formulation of the review question followed the PICo framework introduced by Richardson et al. (1995) specifically for qualitative research, where the population/patient/problem (P) was women, the interest (I) related to a defined event, activity, experience, or process was effective patient-provider communication, the context (C) as in settings was antenatal care in England, and the outcome (o) was experienced (See **Table 1** for PICo worksheet in **Appendix A**). This review sought to find, evaluate, and synthesise all types of observational studies, surveys, qualitative systematic reviews, and thematic analyses that assessed women’s experiences regarding effective patient-practitioner communication in prenatal settings across England. Search strategies were devised and run between September and November 2023 on the following electronic databases to identify relevance: PubMed (from 2010) and The Cochrane Library (from 2010). This timeframe was purposely selected to allow for a comprehensive assessment of women’s experiences following strategies implemented post-2010. Considering the complex nature of assessing women views, which typically evolve gradually and resist immediate shifts, an extended timeframe was deemed necessary to capture extensive data. The search protocol employed key terms used in consistently formulated queries, utilising Boolean operators ‘AND’ across columns and ‘OR’ within columns to retrieve relevant studies during the search process (see **Table 2**).

**Table 2.** Search terms. *Table illustrates key terms generated and used for a literature search strategy*.

| **Population/patient/problem** | **Interest** | **Context** | **Outcome** |
| --- | --- | --- | --- |
| Women | Effective communication | UK | Experience |
| Pregnant | Patient-provider communication | United Kingdom | Perception |
| Patient |  | England | Perspective |
| Mother | Communication | Great Britain | Attitude |
| Maternal | Barriers to communication | Antenatal care | Insight |
| People |  | Prenatal care | View |
| Healthcare professionals | Communication difficulties | Maternity care | Opinion |
| Healthcare providers |  | Maternity services | Barriers |
| Midwife |  |  | Difficulties |
| Consultant |  |  | Satisfaction |
| Obstetrician |  |  |  |

### Eligibility Criteria

The following search strategy generated a reasonable record of articles. Nevertheless, a notable shortage of current research on the topic was identified; therefore, to assist a comprehensive exploration, the analysis included studies conducted from 2010 onwards, allowing for a broader exploration of the subject matter. Studies were screened through independent titles and abstract screening using pre-defined inclusion criteria (see **Table 3** for inclusion and exclusion criteria). Additionally, a reference list of all included studies was hand-searched. Citations were downloaded into the ENDNOTE library, and following this, all duplicates were removed. The studies that met the criteria were fully obtained and assessed to determine inclusion.

**Table 3.**
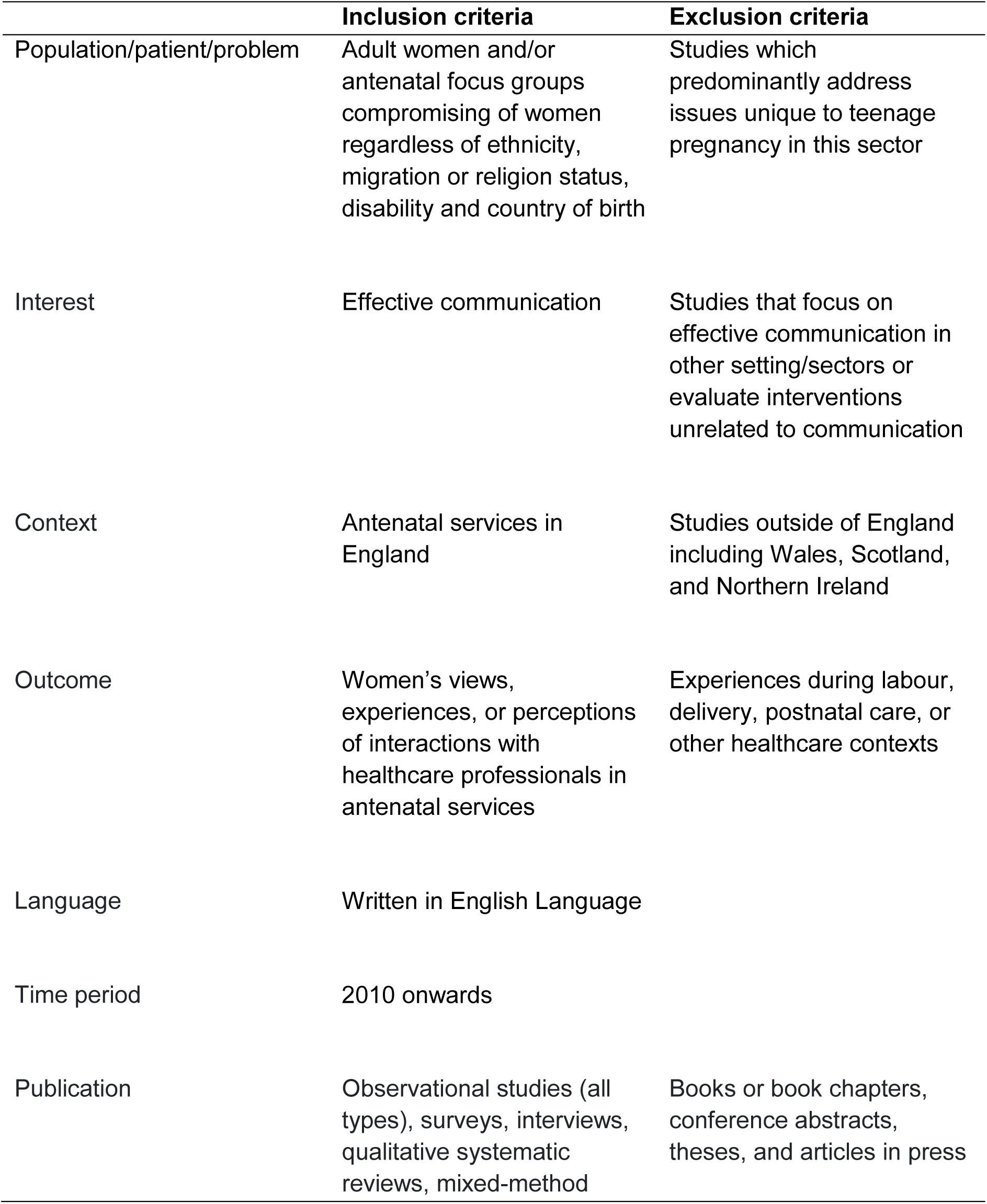
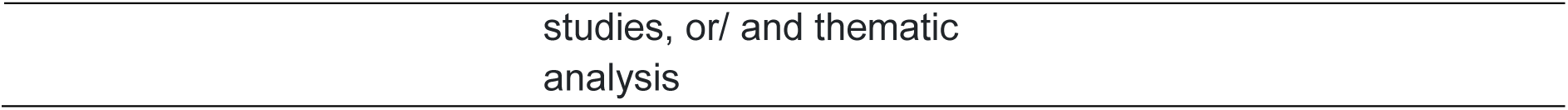
Inclusion and exclusion criteria. *Table illustrates pre-defined inclusion and exclusion criteria for study screening and the assessment of eligibility*.

### Critical Appraisal and Risk of Bias Assessment in Individual Studies

In commencing the examination of relevant qualitative research articles and to provide an in-depth understanding of women’s perspectives and experiences of prenatal care in England, this study aimed to thoroughly synthesise the findings, reliability, and potential limitations inherent in each study considered. Considering that the synthesis of qualitative research is still an expanding and evolving methodological area, to understand strengths, weaknesses, and potential for bias, the Joanna Briggs Institute (JBI) for systematic reviews and research syntheses (2017) tool developed by Pearson et al. (2005) and updated by Lockwood et al. (2015) was used to assess each selected article (See **Table 4** for a summary of critical appraisal using the JBI checklist in **Appendix A-D**). Moreover, according to Porritt et al. (2014), the JBI tool is more appropriate for evaluating qualitative studies and more sensitive to aspects of validity than the critical appraisal skills program (CASP).

### Approach to Analysis

All relevant studies were read thoroughly, and the findings were summarised, acknowledging key conclusions about women’s experiences related to antenatal care in England. Sample size, year of study and study size (i.e., qualitative interview study, qualitative systematic review, population-based survey) were also noted. Study findings were recorded for ease of interpretation and discussion. To extract the data using the qualitative synthesis method, the study used a qualitative descriptive approach, which is a favoured method when a straight portrayal of the phenomena is needed at the initial stage, to obtain the maximum difference of experience and extent of perception prior to consequent thematical evaluation. In conclusion, descriptive thematic analysis of the data obtained was commenced by developing codes for portraying findings, which were later reviewed in relation to coded extracts of entire datasets and summarised with supporting quotes and a narrative that addressed the research question.

## Results

### Study Selection

A comprehensive search across digital databases and supplementary approaches yielded 46 distinctive study reports. Among these, 23 were entirely procured, and six fulfilled the predetermined inclusion criteria upon examination (see **Figure 2**). Notably, one study meeting the criteria involved 13 male participants; however, it was still considered for inclusion due to the significant number of women (385) in the overall dataset, outnumbering the male participants (385:13). Similarity, one of the reviews included within this study included a total of 128 women, 17 HCPs and nine interpreters. Though, this was not considered for the review, and only women’s experiences were evaluated. Inclusive considerations within this study were also extended to studies where there was no explicit assertion to influence on effective communication but where there was at least a part of the research. Nonetheless, all selected studies equally focused on exploring women’s viewpoints and experiences regarding ANC. Additionally, one study provided additional insights into the experiences and insights of women through post-delivery care (Puthussery, 2010), but it was deemed beyond the scope of this review.

**Figure 2.**
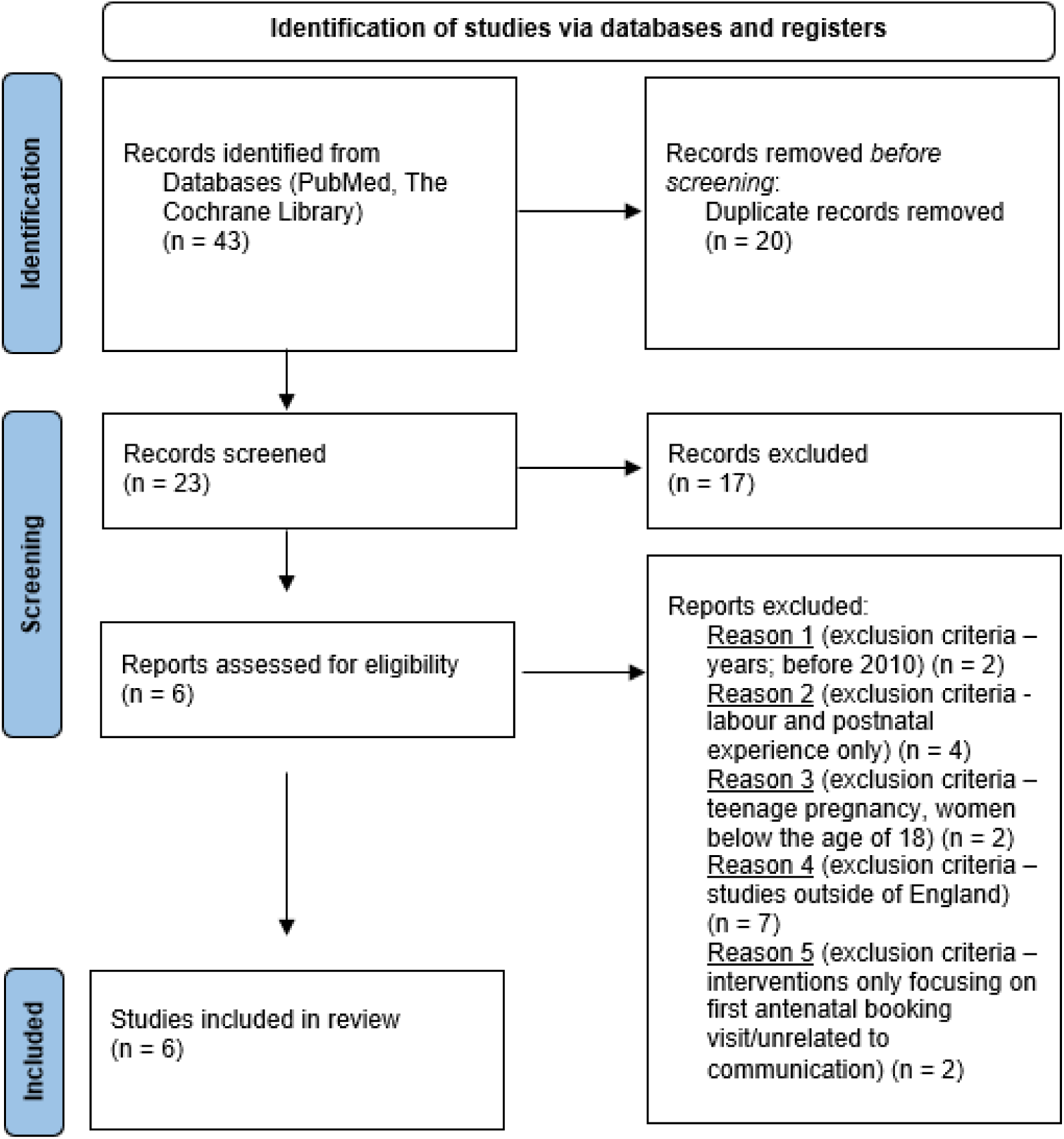
PRISMA flow diagram. Figure utilises PRISMA (2020) flow diagram for new systematic reviews which includes searches of databases and registers, with accordance to the updated guideline for reporting systematic reviews by Page et al., (2021).

### Study Characteristics

Two studies employed a mixture of focus group discussions and semi-structured interviews (Raine et al., 2010 & Thomson et al., 2013); one utilised solely semi-structured interviews (Puthussery et al., 2010); another was a qualitative systematic review, which evaluated six studies for analysis (Firdous et al., 2020), while the remaining two employed online surveys, community consultations, follow-up interviews (Thomson et al., 2022), and narrative semi-structured telephone and Skype interviews (Collins et al., 2023) (See **Table 6** for a summary of the included studies). Two studies specifically delved into the perspectives of ethnic minority women in England (Puthussery et al., 2010 & Thomson et al., 2022), with one focusing on Muslim women (Firdous et al., 2020) and the other on women with physical or sensory disabilities (Collins et al., 2023). Given that two studies focused on minority groups and one focused on Muslim women, one might consider it inadequate; however, as the aim of the research is to delve into the experiences of a diverse range of women, the findings remain applicable to all women, particularly given the era of super-diversity and the expanding multi-faith society in England (Grillo, 2010; Vertovac, 2019). This assertion is reinforced by the fact that one in four births in England and Wales in 2016 was to foreign-born women, which accounted for 28.4% of all births, and this statistic represented the highest level on record (ONS, 2017). This figure has been increasing and, as of 2022, already accounts for 30.3% of all births (ONS, 2023). Moreover, including a study representing disabled women (n = 10) (Collins et al., 2023) adds a valuable perspective. However, most of its findings align with those applicable to all women, irrespective of disability.

**Table 6.**
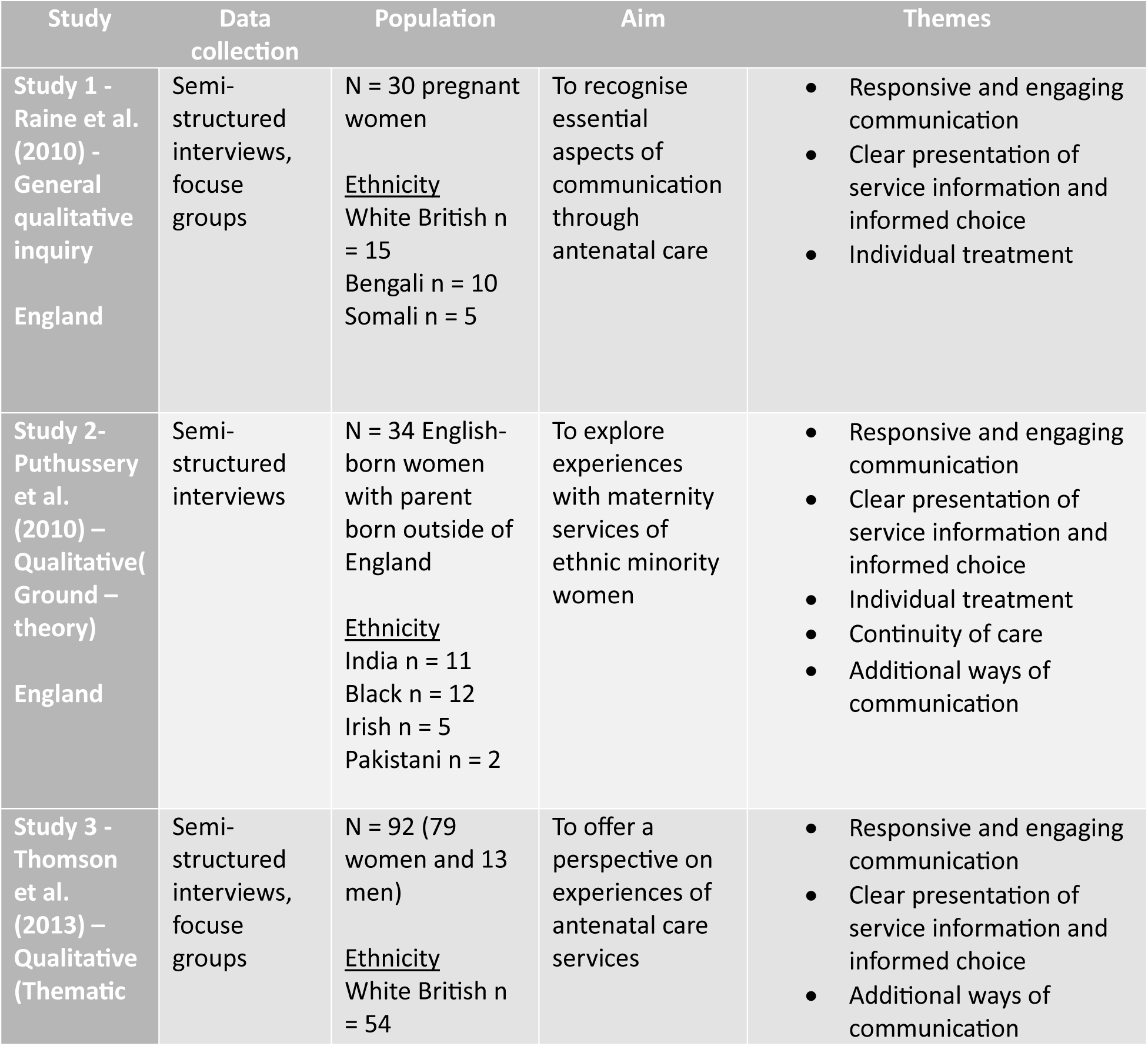
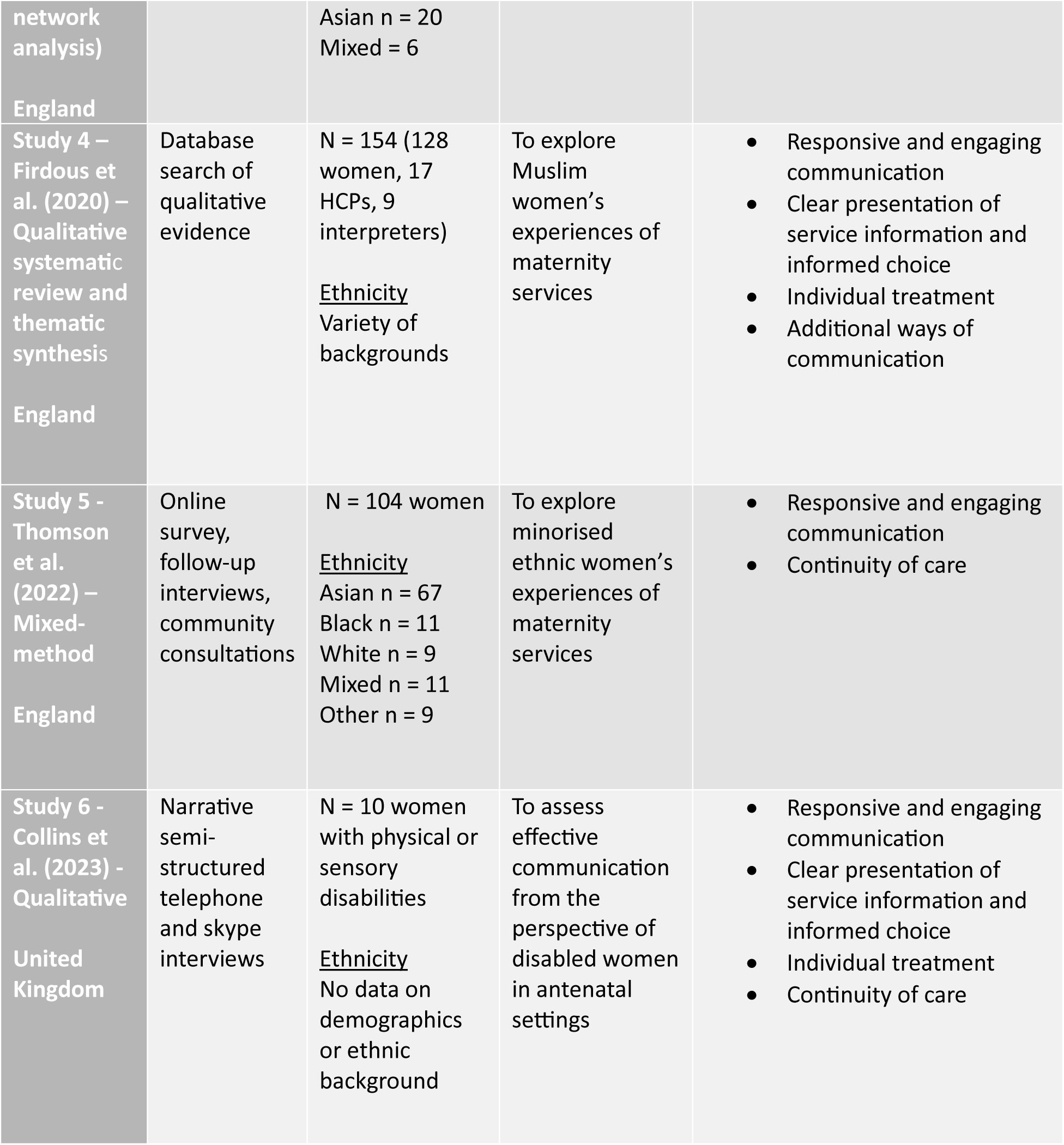
Study characteristics and specifications. *Table illustrates an overview of the included studies, summarising their key characteristics and specifications*.

### Data Extraction

To combine different qualitative research, a thematic synthesis approach developed by Thomas and Harden (Thomas & Harden, 2008) was followed to allow for further interpretation and summarisation of the studies involved. This methodology aided the interpretation and synthesis of the included studies. All text within the result and findings sections of the selected studies, including participant quotations and interpretations by the study authors, was considered as data. Per the framework defined by Thomas and Harden, the codes were subsequently arranged into themes for description. Certain themes exhibited similarities with those observed in the original studies. However, integrating the studies involved allowed for a much broader identification of themes and therefore, significant themes that arose from the data include:

#### Theme 1: Responsive and engaging communication

**Figure 3.**
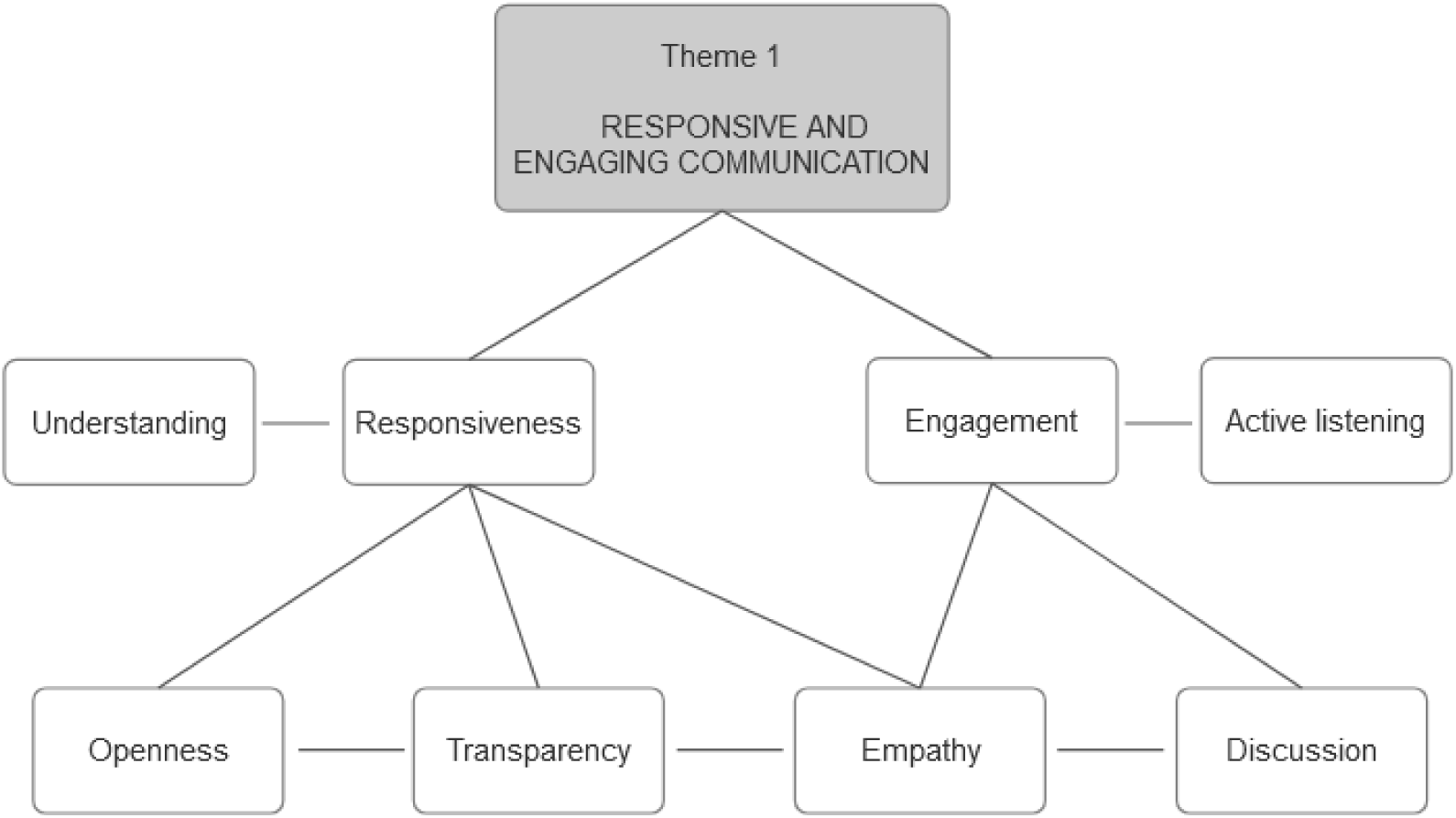
Classification tree of theme (1) with associated codes. Figure illustrates summary of a theme and codes arising from studies involved within the review. Created with BioRender.com.

This theme explores women’s experiences of effective patient-provider communication as open, transparent, responsive, dynamic, and emotionally engaging. Such concepts were running through all six studies involved in this review. The significance of communication emerged in each narrative, with women reporting positive encounters with healthcare providers who demonstrated empathy, actively listened, and responded with understanding and support, similarly reporting negative experiences with staff who did not respect their dignity and rights, did not provide enough attention and time to listen to women concerns about their circumstances and hopes for the pregnancy, leaving many women feeling ignored or even feeling as they had to initiate communication on their own.

> *,, There were so many occasions where I had to chase or trace up.’’ (participant quotation, Study 1, p. 596)*
>
> *,, I tried to tell them. I felt like they’re not hearing me.’’ (participant quotation, Study 1, p. 595)*
>
> *,, She didn’t fob me off, she came around straight away and she really listened to me. … She gave me that time, because she was listening to me… actually feeling that someone is listening to you… that makes a massive difference.’’ (participant quotation, Study 6, p. 5)*
>
> *,, … I didn’t find her kind, I didn’t think she listened to me and I think those things are important. Not just to women. To everyone. You want to be listened to; you want your health care professional to listen…’’ (participant quotation, Study 6, p. 4)*
>
> *,, They don’t always have the people there for you unless you are high risk’ (participant quotation, Study 3, p. 8)*
>
> *,,Not enough information provided…they give you leaflets and tell you some risks…but I would have like to talk to someone…’’ (participant quotation, Study 3, p. 9)*
>
> *,,I went from easy pregnancy to having anaemia and at that time that I was not educated as to what that could cause…’’ (participant quotation, Study 3, p.9)*

The theme also highlights that women appreciated healthcare providers who were sensitive to their needs and concerns, promoting a trusting and supportive environment. They appreciated the reassuring nature of effective communication, with openness and responsiveness from HCPs contributing to a sense of validation and acknowledgement, making the healthcare experience more compassionate and enjoyable.

#### Theme 2: Individual treatment

**Figure 4.**
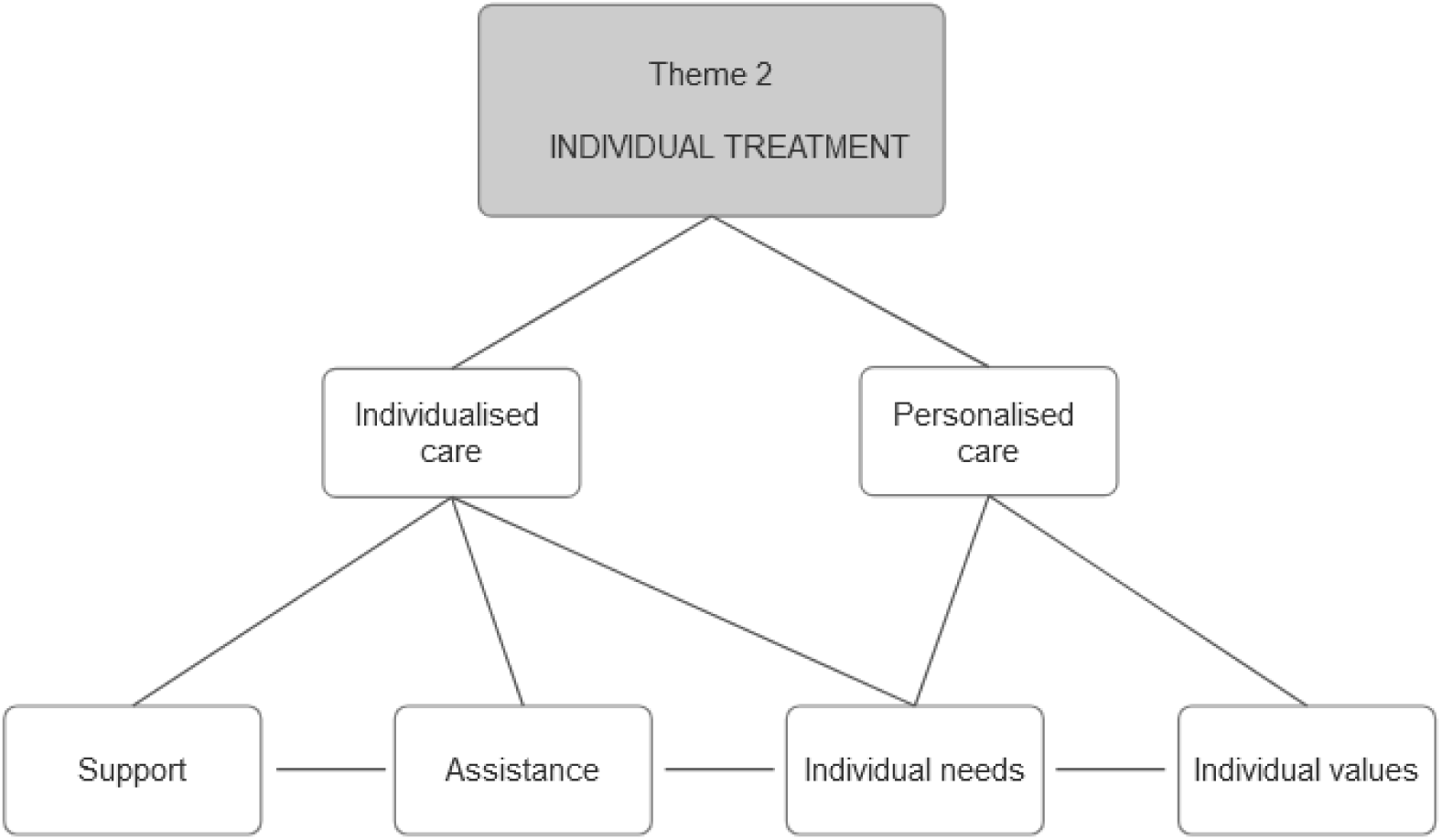
Classification tree of theme (2) with associated codes. Figure illustrates summary of a theme and codes arising from studies involved within the review. Created with BioRender.com.

Individualised treatment was found to play a crucial role in effective patient-provider communication. This theme was evident in four studies and emerged from the perception that healthcare providers should tailor their approach to each woman’s unique circumstances and preferences. This involves personalised care plans, psychological support, consideration of cultural differences, and acknowledging individual needs and values. Participants commonly expressed frustration at the ‘one-size fits all’ approach and a lack of personalisation.

> *,, … It’s a system and it’s a conveyor belt, and you’re just the next body. You’re not a pregnant woman to you’re just a body…’’ (participant quotation, Study 6, p. 6)*
>
> *,, You’re just a number, you’re like an animal. They just don’t care … The service is there, but the care’s not there.’’ (participant quotation, Study 2, p. 158)*
>
> *,, …you’re just another patient and another number really…’’ (participant quotation, Study 2, p. 159)*

Women valued being treated as individuals rather than as a generic patient. This has been especially evident within minority groups, where addressing cultural and social factors was of greater importance. Some women reported that HCPs did not support their cultural and religious meets and that they have experienced racial discrimination within ANC settings. Conversely, UK-born ethnic minority women generally had better experiences with care and less often reported differences in treatment due to their ethnic backgrounds.

> *,, [Did you ever feel that your ethnic background made a difference in the way you were treated?] – No. Absolutely not! Why should I be treated any different from Mary Smith who lives two doors away? … I have access to everything; English is my first language not my second language. Why should I get anything that [is different]? No. In fact I’d resent it if they did! I’d be very annoyed.’’ (participant quotation, Study 2, p. 158)*

Nevertheless, regardless of ethnic background, women emphasised the importance of healthcare providers being attuned and gentle in their communication. Pregnancy was viewed as an emotionally challenging period, during which women perceived themselves as vulnerable, and they anticipated that HCP emphasised their individual needs.

> *,, When they’re dealing with women who are going through the most painful and amazing experience of their life, they need that kind of sense of support and understanding from somebody who’s going to be taking care of them throughout the whole process…’’ (participant quotation, Study 2, p. 158)*

This theme underlines the importance of healthcare providers recognising and respecting the diversity of women’s experiences, especially considering women’s cultural and religious needs, thereby improving the quality and relevance of women-centred and personalised care, which enhances the effectiveness of antenatal care.

#### Theme 3: Clear presentation of service information and informed choice

**Figure 5.**
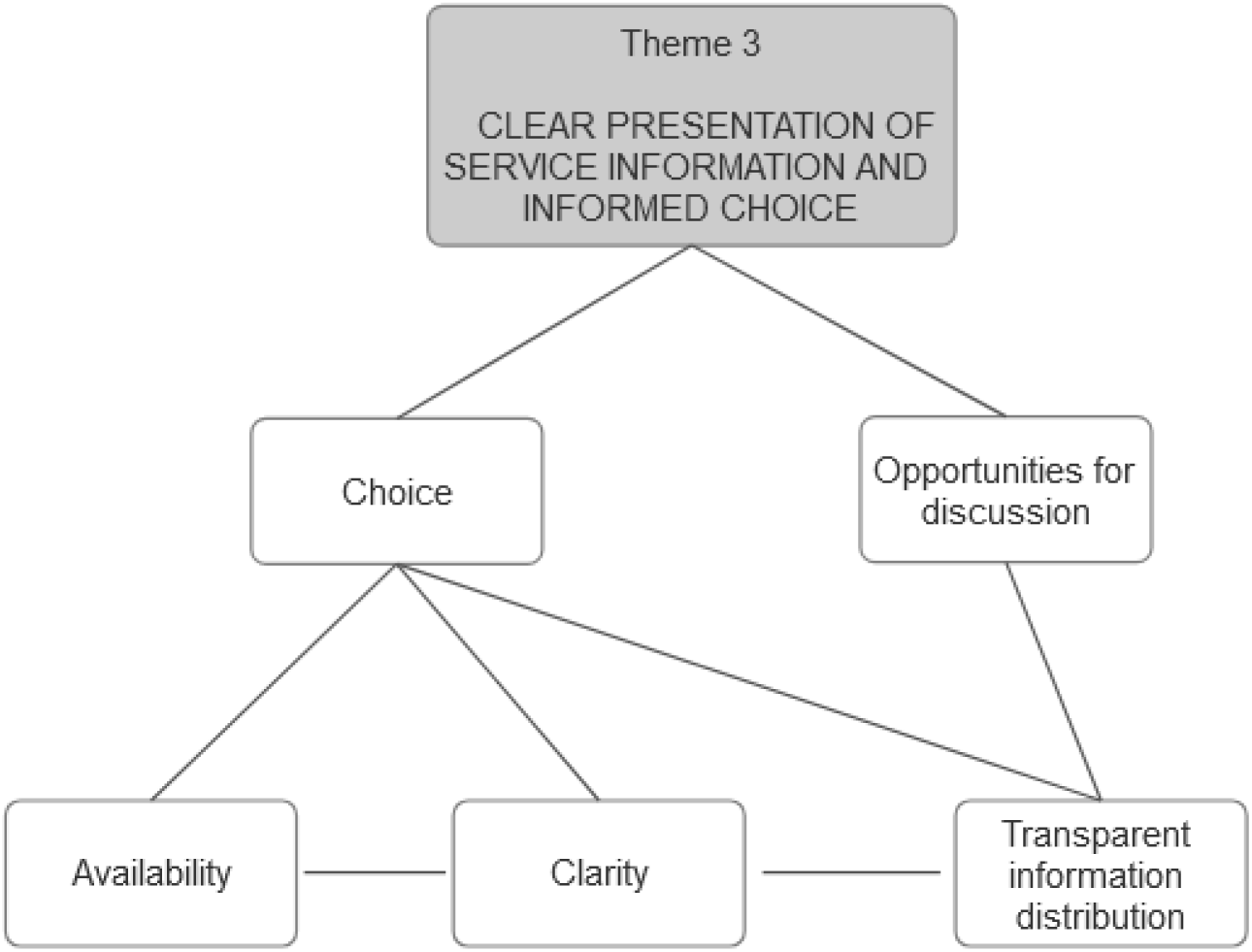
Classification tree of theme (3) with associated codes. Figure illustrates summary of a theme and codes arising from studies involved within the review. Created with BioRender.com.

This theme become apparent in four studies involved and highlights the importance of transparent and easily understandable communication of service information. Women appreciated being well-informed about their options, procedures, and potential outcomes, allowing them to make informed choices regarding antenatal care. Some women experienced a lack of clarity regarding the overall pattern of ANC and felt that HCPs did not consistently explain when specific aspects of care would be addressed, leaving women requiring more information to navigate through the system. Women also felt frustrated with apparent duplications in the purpose of each appointment, which contributed to dissatisfaction.

> *,, It would really help to have a list of numbers saying, you know, this is who you phone for X, this is who you phone for Y… because otherwise you feel lost. ‘’ (participant quotation, Study 1, p. 595)*
>
> *,, They repeat the same thing… ‘’ (participant quotation, Study 1, p. 595)*

Furthermore, women expressed a lack of awareness regarding the roles of different HCPs, with some assuming it was preferable to see an obstetrician even in low-risk pregnancies. This uncertainty extended to understanding certain services or conditions beyond receiving informational leaflets, making it difficult for women to comprehend specific situations and make informed choices fully. It was evident that women also relied on help and support in decision-making. For some high-risk women, there was a perception that specific HCPs lacked knowledge about their conditions, leading to conflicting advice or inadequate guidance. The need to be assertive in requesting suitable care arose, as some specialists and consultants were perceived as dismissive of women’s preferences and opinions in decision-making.

> *,, I’m glad I had confidence in my decisions and that I wasn’t afraid to tell people that was what I wanted.’’ (participant quotation, Study 6, p. 5)*

The theme demonstrated an absence of transparent communication on the overall antenatal pathway, responsibilities of different HCPs, and the purpose of appointments, which left women feeling unsupported and not adequately informed. Clear communication empowered women to contribute actively in decision-making about their healthcare. Feeling informed and involved in the decision-making process enhanced their sense of autonomy and confidence in the care they attained.

#### Theme 4: Continuity of care

**Figure 6.**
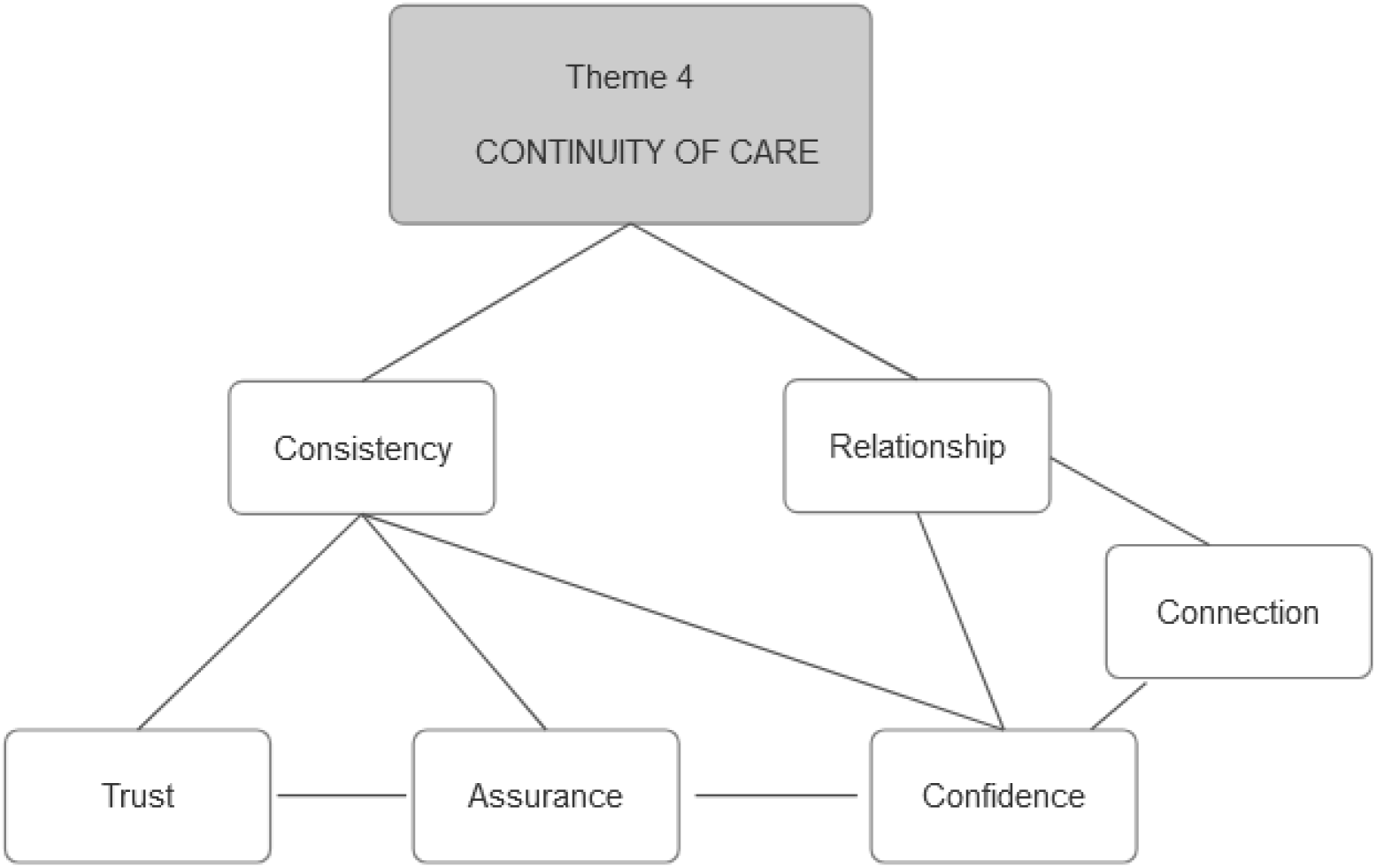
Classification tree of theme (4) with associated codes. Figure illustrates summary of a theme and codes arising from studies involved within the review. Created with BioRender.com.

Evidence from three studies addressed the theme of continuity of care, which implies a continuous and consistent provision of healthcare services where women experience an ongoing relationship with their healthcare providers. This includes consistent follow-ups, which coordinates communication across different stages of the antenatal period. Moreover, women believed that receiving care from an allocated midwife or a consistent team of antenatal care providers mitigates variations in care quality and conflicting advice during pregnancy. They have also reported continuity of care as an essential factor enabling good communication. This preference aligns with effective patient-provider communication, emphasising the importance of developing a trusting and continuous relationship. Women felt they could develop trust and confidence by having the same midwives throughout their care. Changing HCPs, as reported by some women, introduces confusion.

> *,, One thing I always say is I would’ve preferred it if I had a, like a consistent person to see… because you get so many, it was quite confusing…during my pregnancy, I must have seen about ten, fifteen midwives.’’ (participant quotation, Study 2, p. 159)*
>
> *,, It was hard seeing a different obstetrician every single time. I think I saw 4 different obstetricians…’’ (participant quotation, Study 6, p. 6)*
>
> *,, I mean it wasn’t a bad relationship but no continuity, it’s just the fact that you are with different people at every appointment … you don’t really build up a relationship with any of them’’ (participant quotation, Study 2, p. 159)*

Women felt secure and comfortable when care was continuous and consistent. Knowing their healthcare providers and having a consistent point of contact contributed to a more comprehensive and integrated care experience, positively influencing their overall satisfaction, and highlighting the impact of consistent communication on the overall antenatal care experience.

#### Theme 5: Additional ways of communication

**Figure 7.**
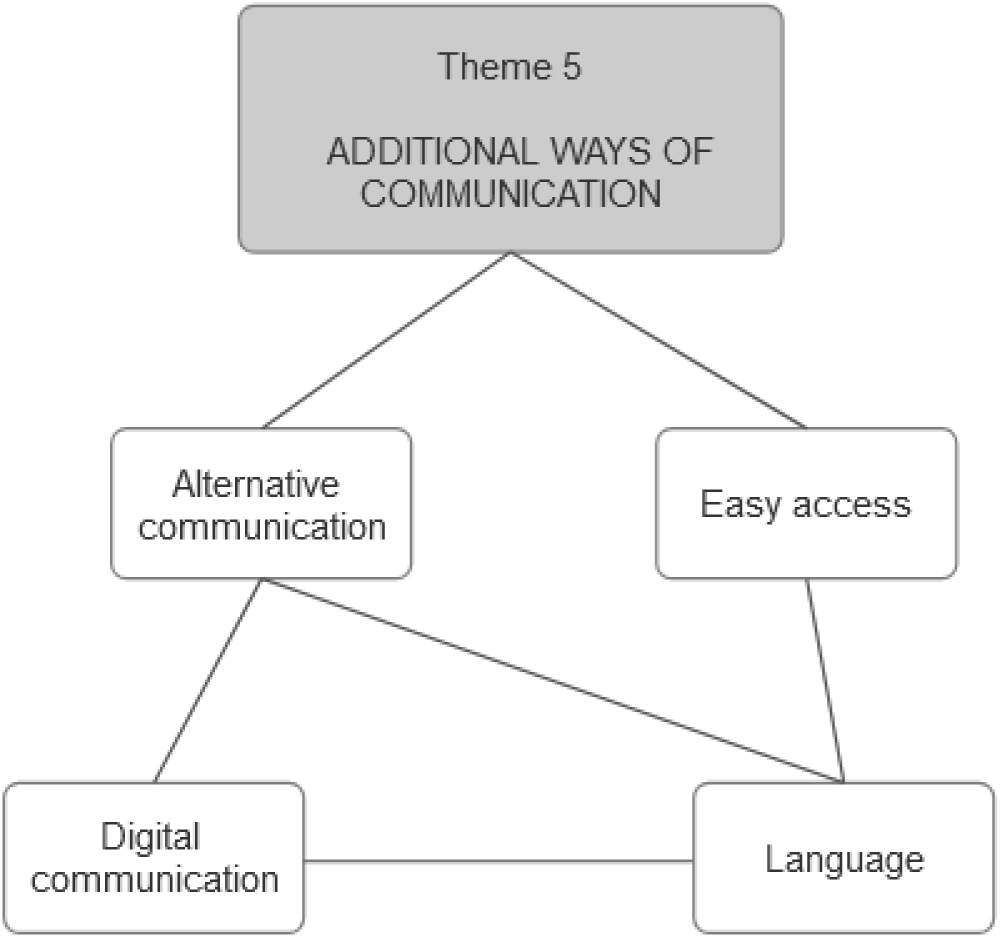
Classification tree of theme (5) with associated codes. Figure illustrates summary of a theme and codes arising from studies involved within the review. Created with BioRender.com.

This theme incorporates the use of alternative communication channels beyond traditional face-to-face interactions and was also evident within three studies. Women valued the accessibility of additional non-verbal communication methods. These alternative channels assisted with easier access to information, contributing to a more connected and efficient healthcare experience. Examples include text messages, online platforms, or other forms of digital communication that complement and enhance the overall patient-provider communication experience. Women appreciated additional ways of communicating with their designated HCP through messages, exchanging information about results, or reminders for future appointments, and general pregnancy related concerns.

> *,,I think you should have a point of access to see someone when you are worried, to have that flexibility with the service rather than a set of points.’’ (participant quotation, Study 3, p. 8)*

Evidence from study number 4 has also highlighted language barriers and the need for additional ways of communication in this sector, which can only be achieved with proper translation. Women expressed challenges in accessing maternity services, citing a limited choice of interpreters. Consequently, women resorted to using a family member or friend as interpreters.

> *,,… I could not understand everything they said. I told my husband to translate everything for me, but he did not. He was hiding the truth and trying to comfort me.’’ (participant quotation, Study 4, p. 11)*

## Discussion

This systematic review studied women’s experiences of effective patient-provider communication within antenatal services in England. It identified five key themes that substantially contribute to constructive communication and offer insight into the complex aspects of women’s interactions with their caregivers. The key finding stressed the fundamental role of emotional connection and empathy. Women’s narratives consistently emphasised the positive impact of HCPs, who actively listened, demonstrated sympathy, and responded with understanding and support. These discoveries are supported by Goberna-Tricas et al. (2011) and Novick (2009), who also previously reported that women want caring, reassuring, and engaging providers. Likewise, the study found that responsive and engaging communication facilitated a constructive relationship with HCPs, which, more than everything else, allowed effective information exchange, promoting fruitful interactions with caregivers. Furthermore, women expressed that effective communication predominantly involved being heard by medical professionals. Above all else, it allowed or inhibited the establishment of trust, which was crucial in creating the desired connection through communications. This feeling generally influenced women’s overall satisfaction with ANC, and the lack of these features contributed to difficulties related to the overall care experience. Women have reported similar themes in the 2016 UK National Maternity Review, Better Births (National Maternity Review, 2016), which highlighted a need for maternity services to become kinder and more professional, amongst other recommendations.

Other significant findings that enhanced communication effectiveness in the researched sector revolved around the concept of individualised treatment and a desire to feel more than just a number, with participants in this review expressing frustration with a perceived ‘one-size-fits-all’ approach. This critical aspect points to the importance of adapting and shaping care approaches to each woman’s unique circumstances and preferences, who empathised with the need to discuss each of their personal needs openly. Women also underlined the importance of cultural and religious sensitivity and general support at any stage of ANC. This information mirrors WHO recommendations, which identified a women-centred, personalised care approach as a priority for safe, high-quality care (WHO, 2016). Moreover, results from this study have been echoed internationally, as demonstrated by Bohren et al. (2020), who highlighted individualised treatment as an important aspect of respectful maternity care. Additionally, continuity of care emerged as a crucial factor influencing effective communication, often highlighting a preference for maintaining a consistent relationship with HCPs throughout the prenatal period. The positive impact of continuity of care was marked in women’s experiences, who exhibited more trust and confidence when receiving care from the same midwife or team of providers. In contrast, frequent caregiver changes were found to introduce confusion and disrupt the development of a meaningful patient-provider relationship. The data also emphasises the importance of clear communication about antenatal care options, procedures, and potential outcomes, as crucial factor of interactions. The revealed findings are also supported by Farquhar et al. (2000) and Jenkins et al. (2015), who exclusively investigated women’s perspectives on continuity of care in maternity services and drew similar conclusions about women appreciating smaller midwifery teams, which essentially allows them to see fewer providers during ANC, therefore avoiding unnecessary stress and confusion associated with interactions and communications.

Further insights from this study highlight the need for transparent and easily understandable service information, essential for women to make well-informed decisions concerning antenatal choices. Patterns identified within this review illustrate that women expressed dissatisfaction with unclear communication, leading to frustration and challenges in decision-making. The established theme also illustrates the importance of empowering women through appropriate communication, which helps them actively participate in decision-making about their healthcare. Moreover, evidence uncovered in this study also demonstrates that non-verbal communication methods, such as text messages, contribute to a more connected and efficient healthcare experience. Language barriers have also been identified as a challenge in achieving effective and desired communication with HCPs by some women from other ethical backgrounds, emphasising the importance of appropriate and universal translation services to improve communication and information exchange. These findings were also reflected within a comparative review study of maternity care by Small et al. (2014) in five countries, including the United Kingdom, and found that women often had trouble communicating due to language differences.

Findings therefore align with the hypothesis as they support the perception that effective patient-provider communication significantly influences the emotional well-being and satisfaction of pregnant women during antenatal care. The identified themes highlight the importance of improved communication for promoting trust, confidence, and positive experiences in prenatal care. Moreover, analysis of the key themes produced from this review outlines the critical role of effective patient-provider communication in prenatal care settings, notably with HCPs playing a central role in ensuring the delivery of high-quality antenatal care services, where appropriate communication is embarked upon and delivered. Such an important role requires appropriate skills and a profound understanding of women’s challenges during pregnancy. The findings illustrate the need of HCPs to prioritise building emotional connections and demonstrating empathy as fundamental aspects for positive experiences. Besides, the insights gained from this study provide an understanding of complex communication issues, and the significance of the results becomes apparent when considering guidance for healthcare providers, policymakers, and researchers who seek to enhance the quality of ANC in England. Although suggesting the importance of effective patient-provider communication within the antenatal sector is not new, this study contributes to the evolving understanding of what makes communication effective and how. Nevertheless, despite the recognised importance of antenatal care, a considerable gap still exists in the literature about women’s experiences related explicitly to effective communication between women and their caregivers in England. Apart from National Maternity Surveys, only few studies have tried to verify and synthesise actual women’s experiences. Therefore, this study contributes to filling this void by exploring these experiences and providing insights into what they mean for women.

Additionally, even though the NHS has practically applied its policies and some of its initiatives to improve maternity services, such as the Better Births initiative (National Maternity Review, 2016), more attention must be paid to communication aspects, which lead to persistent and similar issues being identified over the past decade. Moreover, it is worth mentioning that the recent decline in the delivery of ANC could also be marked by the COVID-19 pandemic and its detrimental effects on healthcare (Meaney et al., 2022; Sanders & Blaylock, 2021). However, the recurring identification of concerns through surveys and studies suggests broader systemic issues, such as medical personnel often being constrained by barriers of time, training, and resources, as well as staff burnout and low morale (Cooksley et al., 2023). Initiatives to enhance ANC services must be addressed by applying and evaluating policies related to effective communication. This calls for more action from commissioners to improve the outcomes of health inequalities and highlights the need for structural changes in the healthcare sector, mainly focusing on comprehensive communication and cultural safety training for HCPs. Future interventions should target issues related to communication in more detail; training programmes for HCPs could focus on active listening, sympathy and understanding, as well as promoting individualised care, continuity of care, and transparent distribution of information through various communication channels, as despite guidance about communication in ANC settings, evidence suggests that not all women receive appropriate care. Moreover, efforts should be made to empower women through appropriate communication, encouraging their active and enthusiastic involvement in decision-making about their healthcare. This study also reveals a functional gap in prenatal services, highlighting the urgency of addressing communication issues for a more positive antenatal care experience for all women regardless of their ethnicity or backgrounds. Challenges associated with issues in this sector should also be evaluated against other external barriers to maternal health services, such as prolonged underinvestment (Sumankuuro et al., 2018) and staffing shortages (Edwards et al., 2020; Sandall et al., 2011). This study forms the basis for future interventions and research in this critical healthcare sector.

### Strengths and limitations

The social diversity of the sample involved defines the primary advantage of this study. By including women from various social and ethical backgrounds, this study investigated patterns and challenges in patient-provider communication that might otherwise be overlooked in a more homogenous sample. Diversity also assists in a more comprehensive understanding of the topic. It ensures cross-cultural validity, especially in England, where the country’s multicultural and diverse population influences the importance of considering women from various backgrounds. However, social diversity and consideration of minority ethnic women may pose certain limitations as women from these backgrounds are at greater risk of mistreatment and can, more often, report issues related to discrimination and may also experience worse outcomes from their antenatal care (Sharma et al., 2023). Therefore, findings may not have generated sufficient depiction of the experience of women in general population in England. Additionally, it is crucial to acknowledge the inherent challenges associated with measuring women’s views and perceptions of effective patient-provider communication during pregnancy. The emotional intensity, occasional high levels of stress, and the physical demands of pregnancy create a context where the measurement of experiences becomes particularly complex. Despite this complexity, the review identified notable themes, indicating important similarities in women’s experiences.

Moreover, the timeframe covered by the studies included in this review, spanning from 2010 to 2023 (2010, 2010, 2013, 2020, 2022, 2023, respectively), enhances the study’s strength by providing a broader understanding of issues across the years. While the selected studies do not exclusively focus on effective communication, they offer insights into the challenging nature of patient-provider interactions within the ANC setting. However, it is essential to acknowledge that this breadth may pose some limitations regarding the depth of data related to effective communication. Despite this limitation, the review effectively identifies similarities and trends across the examined studies, contributing valuable insights to understanding patient-provider communication in the context of antenatal care in England.

## Conclusion

The study’s identified themes highlight the fundamental role of effective patient-provider communication in prenatal settings in England. The strategies proposed, including continuity of care, clear service information, engaging communication, individualised treatment, and additional ways of communication, emphasise practical steps to enhance women’s experiences. However, addressing structural issues within the NHS is crucial for successful implementation, which requires ensuring that government initiatives align with identified themes and actively address any systemic barriers. The study calls for further research to explore emerging challenges, particularly in existing circumstances, to continually inform evidence-based practices and policies, optimising and improving patient-provider communication in ANC settings, for the well-being of mothers and their children in England.

## Data Availability

All data produced in the present study are available upon reasonable request to the authors

