## Appendix for "Examining Effective Patient-Provider Communication in Antenatal Settings across England: An In-Depth Analysis of Woman’s Experiences"

**Appendix A**

**Table 1. PICo worksheet**

*Table illustrates the most important components of research question as defined by PICo framework for qualitative research introduced by Richardson et al. (1995).*

| Population/Patient/Problem - What are the characteristics of the **P**opulation or **P**atient? What is the **P**roblem, condition or disease you are interested in? | Pregnant women |
| --- | --- |
| Interest - **I**nterest relates to a defined event, activity, experience, or process. | Effective patient-practitioner communication |
| Context - **Co**ntext is the setting or distinct characteristics | Antenatal Care in England |
| Research question | What are the experiences of women regarding effective patient-practitioner communication in antenatal care services across England? |

**Table 4. Summary of critical appraisal using the JBI checklist**

*Table illustrates Joanna Briggs Institute JBI (2017) Critical Appraisal Checklist for Systematic Reviews and Research Syntheses used to assess included studies produced by Lockwood et al. (2015).*

Author: Raine et al. Year: 2010	Study Number: 1

1. Is there congruity between the stated philosophical perspective and the research methodology? Yes

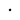

2. Is there congruity between the research methodology and the research question or objectives? Yes

3. Is there congruity between the research methodology and the methods used to collect data? Unclear

4. Is there congruity between the research methodology and the representation and analysis of data? Yes

5. Is there congruity between the research methodology and the interpretation of results? Yes

6. Is there a statement locating the researcher culturally or theoretically? No

7. Is the influence of the researcher on the research, and vice- versa, addressed? Yes

8. Are participants, and their voices, adequately represented? Yes

**Appendix B**

9. Is the research ethical according to current criteria or, for recent studies, and is there evidence of ethical approval by an appropriate body? Yes

10. Do the conclusions drawn in the research report flow from the analysis, or interpretation, of the data? Yes

Overall appraisal: Include

Comments: The broader literature context was considered in the discussion of the findings, with an exploration of transferability and the identification of a new research area.

Author: Puthussery et al. Year: 2010	Study Number: 2

1. Is there congruity between the stated philosophical perspective and the research methodology? Yes

2. Is there congruity between the research methodology and the research question or objectives? Yes

3. Is there congruity between the research methodology and the methods used to collect data? Yes

4. Is there congruity between the research methodology and the representation and analysis of data? Unclear

5. Is there congruity between the research methodology and the interpretation of results? Yes

6. Is there a statement locating the researcher culturally or theoretically? No

7. Is the influence of the researcher on the research, and vice- versa, addressed? Yes

8. Are participants, and their voices, adequately represented? Yes

9. Is the research ethical according to current criteria or, for recent studies, and is there evidence of ethical approval by an appropriate body? Yes

10. Do the conclusions drawn in the research report flow from the analysis, or interpretation, of the data? Unclear

Overall appraisal: Include

Comments: The data analysis lacked rigor, and minor concerns were identified with explicit statements related to the findings. Additionally, no new research area was identified.

Author: Thomson et al. Year: 2013	Study Number: 3

1. Is there congruity between the stated philosophical perspective and the research methodology? Yes

2. Is there congruity between the research methodology and the research question or objectives? Yes

3. Is there congruity between the research methodology and the methods used to collect data? Unclear

4. Is there congruity between the research methodology and the representation and analysis of data? Yes

**Appendix C**

5. Is there congruity between the research methodology and the interpretation of results? Yes

6. Is there a statement locating the researcher culturally or theoretically? No

7. Is the influence of the researcher on the research, and vice- versa, addressed? Yes

8. Are participants, and their voices, adequately represented? Yes

9. Is the research ethical according to current criteria or, for recent studies, and is there evidence of ethical approval by an appropriate body? Yes

10. Do the conclusions drawn in the research report flow from the analysis, or interpretation, of the data? Yes

Overall appraisal: Include

Comments: Exclusion criteria was not mentioned, However a thorough examination of the findings was conducted within the broader framework of relevant literature.

Author: Firdous et al. Year: 2020	Study Number: 4

1. Is there congruity between the stated philosophical perspective and the research methodology? Yes

2. Is there congruity between the research methodology and the research question or objectives? Yes

3. Is there congruity between the research methodology and the methods used to collect data? Yes

4. Is there congruity between the research methodology and the representation and analysis of data? Yes

5. Is there congruity between the research methodology and the interpretation of results? Yes

6. Is there a statement locating the researcher culturally or theoretically? No

7. Is the influence of the researcher on the research, and vice- versa, addressed? Yes

8. Are participants, and their voices, adequately represented? Yes

9. Is the research ethical according to current criteria or, for recent studies, and is there evidence of ethical approval by an appropriate body? Yes

10. Do the conclusions drawn in the research report flow from the analysis, or interpretation, of the data? Yes

Overall appraisal: Include

Comments: The data collection generated insights into participant ethnicity and work status. The research conclusions were clear and appropriate.

Author: Thomson et al. Year: 2022	Study Number: 5

1. Is there congruity between the stated philosophical perspective and the research methodology? Yes

2. Is there congruity between the research methodology and the research question or objectives? Yes

**Appendix D**

3. Is there congruity between the research methodology and the methods used to collect data? Yes

4. Is there congruity between the research methodology and the representation and analysis of data? Yes

5. Is there congruity between the research methodology and the interpretation of results? Yes

6. Is there a statement locating the researcher culturally or theoretically? No

7. Is the influence of the researcher on the research, and vice- versa, addressed? Yes

8. Are participants, and their voices, adequately represented? Yes

9. Is the research ethical according to current criteria or, for recent studies, and is there evidence of ethical approval by an appropriate body? Yes

10. Do the conclusions drawn in the research report flow from the analysis, or interpretation, of the data? Yes

Overall appraisal: Include

Comments: Consistent evidence generated the findings, discussion on the transferability took place.

Author: Collins et al. Year: 2023	Study Number: 6

1. Is there congruity between the stated philosophical perspective and the research methodology?

2. Is there congruity between the research methodology and the research question or objectives?

3. Is there congruity between the research methodology and the methods used to collect data?

4. Is there congruity between the research methodology and the representation and analysis of data?

5. Is there congruity between the research methodology and the interpretation of results?

6. Is there a statement locating the researcher culturally or theoretically?

7. Is the influence of the researcher on the research, and vice- versa, addressed?

8. Are participants, and their voices, adequately represented?

9. Is the research ethical according to current criteria or, for recent studies, and is there evidence of ethical approval by an appropriate body?

10. Do the conclusions drawn in the research report flow from the analysis, or interpretation, of the data?

Overall appraisal: Include

Comments: Minor data collection concerns arose, as the study population's ethnicity was omitted for confidentiality. Study findings were clear and directly descriptive, new are of research was identified.
